# A Scalable Method for Validated Data Extraction from Electronic Health Records with Large Language Models

**DOI:** 10.1101/2025.02.25.25322898

**Authors:** Timothy J. Stuhlmiller, AJ Rabe, Jeff Rapp, Alaa Awawda, Hiba Kouser, Kristi Lui, Hugh Salamon, Donald Chuyka, William Mahoney, John M. Furgason, Madhuri Paul, Frank J. Scarpa, Santosh Kesari, Mika Newton, Kenny K. Wong, Glenn A. Kramer, Mark A. Shapiro

## Abstract

**Purpose/Background:** Healthcare organizations increasingly require structured, patient-level clinical variables for treatment decisions, operational workflows, quality measurement, and clinical trial screening. Relevant information is often fragmented across heterogeneous EHR systems, unstructured formats, and scanned documents. Large language models (LLMs) offer an opportunity to enhance medical record data extraction, particularly in oncology where point-of-care structured coding is often insufficient to capture a patient’s longitudinal clinical course.

**Methods:** Two complementary LLM-based approaches were developed. In the first, LLMs performed schema-based named entity recognition and relation extraction from unstructured documents; extracted variables were normalized to FHIR and OHDSI vocabularies. In the second, a retrieval-augmented checklist framework queried and integrated embedded document text with extracted structured data using prescriptive prompts to answer specific clinical questions, returning custom outputs with evidence-based justifications and source-document citations. Performance was evaluated through human validation, automated consistency checks, and iterative error analysis.

**Results:** Schema-based extractors designed to extract medications, radiation, and surgical procedure fields achieved∼95% accuracy, precision, recall, and F1. Deployed across 3,493 patients, LLM extraction yielded 71% more total medication records and a 207% increase in distinct oncology drug ingredients over structured C-CDA sources. Oncology therapies were captured for 65% of patients versus 40% in structured data, with dramatically improved clinical attribute coverage: indication for prescription (70.5% vs. 4.6%) and reason for discontinuation (9.7% vs. 0%). For radiation and surgical procedures — largely absent from structured records — LLM extraction yielded a 715% and 422% increase, respectively.

The checklist extraction framework achieved significant F1 scores when designed to extract cancer diagnosis variables (99.0%) and lines of therapy (97.6%) across 4,802 validated elements. Cancer diagnoses and dates were identified for 93.5% of patients versus 69.8% in structured data; stage and grade were extracted for 64% and 62% of patients versus near-zero structured availability. A lines-of-therapy checklist generated 4,218 treatment lines across 2,320 patients, capturing regimen dates, best response, discontinuation reasons, and progression dates. Among patients with response data, objective response rate declined from 64.7% (95% CI 59.0–70.0%) at first line to 21.5% (95% CI 13.3–33.0%) at third line, and first-line response rates varied across tumor types from 42.9% in colorectal to 85.0% in esophageal cancer.

**Conclusions:** Leveraging two complementary LLM-based strategies, we substantially enhanced the completeness and utility of structured clinical data from heterogeneous medical records, supporting scalable generation of interoperable patient-level datasets for clinical analytics, research, and operational workflows.

## Introduction

Healthcare data interoperability has increased the sharing of clinical information, but due to the diversity of systems and lack of standardized documentation processes, finding key information remains challenging at the point-of-care. AI solutions may help clinicians quickly find accurate information, but only if they are able to quickly and accurately parse large amounts of heterogenous records from disparate sources. To this end, we have developed a scalable pipeline to assist clinicians with information retrieval tasks.

Patient medical information is scattered across numerous systems from different practices and institutions. This makes information retrieval challenging even when one can access records across different EHR systems. Furthermore, free text medical notes are designed to facilitate clinical communication, while the structured data in an EHR is designed for billing purposes. Thus, accessing information in notes is essential for continuity of clinical care.

However, clinical notes are semantically and linguistically complex. They include orders which may or may not have been filled, specialized forms of negation, and extensive use of context-dependent abbreviations, which have all proven to be challenging for natural language processing. To overcome this, a wide variety of methods have been tested in academic settings, with rule-based and statistical methods, such as conditional random fields, as well as hybrid and machine-learning methods all able to achieve good performance.^1,2^ However, rules-based systems are difficult to develop and can lack real-world robustness. Machine-learning methods have suffered from limited public datasets upon which to train and the cost and expertise required to create labeled training data.

Notably, those systems fall well short of the 95% accuracy and F1-scores proposed for clinical use following the original I2B2 challenge.^2^ More recently, specialized LLMs have shown strong abilities on these and more complex extraction tasks.^3^ Here, we present a highly scalable solution using general purpose LLMs for accurate clinical text classification and structured data extraction that can be implemented and adapted quickly, achieves targeted performance, enables efficient and effective data retrieval, and integrates with standard healthcare data models.

## Methods

### Core Platform Functions

#### Data Collection and Preprocessing

Patient documents were received in multiple formats, including image files, Fast Healthcare Interoperability Resources (FHIR), and Consolidated Clinical Document Architecture (C-CDA) files. Structured C-CDA data was converted to FHIR R4, preserving tabular structure and coding before running additional coding checks and concept normalization.^4^ Semi-structured and unstructured data, including OCR-processed image files, underwent preprocessing including document parsing, chunking, and metadata extraction before being routed to LLM-based extractors.

#### Embedding Model and Text Chunking

Documents were divided into sections based on C-CDA structure, headings, and content type, with further chunking by paragraph, sentence, or predefined section headings while preserving table structures. A text embedding model generated a vector representation for each chunk. In a sample of 41,900 patient records, an average of 231 documents per patient yielded approximately 335 million embedding vectors for classification and retrieval.

#### Classification and Semantic Search

Document and section labels vary widely across EHR systems, requiring a standardized classification scheme. We used unsupervised clustering to identify an optimal number of document classes, and implemented classification of chunks into 26 predefined information types (e.g., Diagnosis, Medications, Plan of Care, Genomics). A supervised random forest classifier trained on embedding vectors was applied following a standard train-test-validate split, with human review confirming training set accuracy. Micro F1 scores were calculated on a validation set of 21,837 records (Table 1).

**Table 1.** Performance of document classifier machine learning algorithm.

| Classification | F1 score |
| --- | --- |
| allergies_adverse_reactions_alerts | 0.98 |
| assessment_and_plan | 0.98 |
| demographics | 0.92 |
| diagnosis | 0.89 |
| diagnostic_test_or_lab_results | 0.80 |
| disease_biomarkers | 0.98 |
| family_medical_history | 0.98 |
| functional_status | 0.98 |
| genomics | 0.98 |
| history_of_present_illness | 0.97 |
| insurance | 0.99 |
| interventional_radiology | 0.99 |
| medical_equipment_and_implants | 0.99 |
| medications | 0.84 |
| oncology | 0.88 |
| pathology | 0.96 |
| pharmacology_progress | 1.00 |
| physical_exam | 0.96 |
| plan_of_care | 0.97 |
| procedures | 0.94 |
| radiation | 0.99 |
| radiology | 0.91 |
| reason_for_visit | 0.98 |
| social_history | 0.95 |
| surgery | 0.96 |
| vital_signs | 0.96 |

### LLM-based data extraction and summarization

We developed two complementary LLM-based approaches to enhance structured data derived from medical records.

#### Method 1: Bottom-up Schema-Based Extraction and Normalization

This document-centric approach applies LLM-based Named Entity Recognition (NER) and Relation Extraction (RE) to identify and capture verbatim terms and values from unstructured and semi-structured clinical text and convert them into standardized, schema-defined structured data aligned with FHIR R4. The primary objective is to augment pre-structured C-CDA data by capturing elements that are generally absent, incomplete, or inconsistently coded in routine structured feeds — particularly granular oncology variables documented exclusively in narrative reports, such as staging, histology, procedure parameters, and molecular results. This schema-based approach systematically captures and normalizes strings to augment the structured patient data, complete with anchored document provenance.

#### Method 2: Top-down Checklist-Based Patient-Level Assertion with Automated Semantic Retrieval

This question-centric approach generates patient-level clinical determinations across the longitudinal record by applying prescriptive LLM prompts to answer specific clinical questions, returning structured outputs (boolean, categorical, date, numeric, or array) with evidence-based justifications and source-document citations. A Retrieval-Augmented Generation (RAG) workflow automatically identifies candidate documents via semantic search, refined by checklist-specific filters (document classification, temporal windows).^5,6^ Prompts explicitly encode evidence hierarchy rules for adjudicating conflicting documentation — for example, prioritizing pathology reports over clinic notes for cancer staging — to assert a single, clinically accurate patient-level conclusion from the available evidence.

#### Interaction Between the Two Methods

The two strategies are complementary: schema-based extraction increases the breadth and density of structured data by harvesting granular events from narrative text and scanned reports, while checklist-based assertion increases clinical correctness and usability by performing patient-level adjudication across conflicting sources to produce a single, evidence-grounded clinical conclusion.

#### Document and Schema Selection Logic for Extraction

Documents and subsections are matched to extraction schemas via semantic search using schema-specific terms, followed by document classification. For C-CDA documents, section headers and the random forest classifier determine schema assignment; for OCR documents, the classifier alone is used. Schemas are designed around focused, related concept sets to optimize LLM accuracy accuracy and consistency while minimizing hallucinations.

#### Prompt Engineering and Model Refinement

Accurate LLM extraction is achievable with well-designed zero-shot prompts but improves with in-context learning using manually annotated and synthetic examples.^7^ Extraction errors — whether from retrieval mismatches or prompt-level failures — are captured systematically and used to iteratively refine prompts, retrieval parameters, and dataset augmentation. A/B testing across prompts, models, and hyperparameters guides ongoing refinement.

### Standardization

#### Mapping to FHIR

C-CDA are initially passed through a pipeline to generate pre-structured medication data and medical document metadata, mapped to FHIR R4. Schema-based LLM extraction outputs are also mapped to FHIR R4 resources for interoperability whenever possible. Data not covered by a FHIR mapping is stored in extension fields per published profiles (e.g., mCODE), or a custom data model where no suitable profile exists.^8^

#### OHDSI Vocabulary Normalization

After extraction, further processing is required to integrate these concepts with the broader medical record. FHIR resources mapped from C-CDA contain a code from an Observational Health Data Sciences and Informatics (OHDSI) Standardized Vocabulary System (e.g., LOINC or SNOMED).^9^ LLM extractions and a subset of C-CDA-derived data lacking OHDSI vocabulary mappings are matched to high-confidence codes, with manual mapping applied where automated matching fails. Normalized concepts are grouped into high-level clinical categories (e.g., “immunotherapy,” “cytotoxic chemotherapy”) using key nodes in medical ontologies to support downstream analytics.

### Validation

Clinically-trained human reviewers independently assessed extraction outputs against source medical documents, with a random 10% audit for reliability and discrepancies refereed by a third reviewer with access to clinical experts.

For schema-based extractions, reviewers validated that extracted values were verbatim as documented. Hallucinations in this context typically represent correct inferences not explicitly stated in source text (e.g., inferring route “PO” from “APAP 325mg”) — a failure of faithfulness rather than factualness.^10,11^ Only explicitly stated, verifiable extractions were counted as True Positives.

For checklist-based assessments, a two-stage adjudication process was used. Reviewers first assessed extracted outputs against cited source documents, classifying results as TP or FP. They then conducted an independent semantic search of the full EHR to identify contradicting evidence (confirming FP) or relevant missed evidence (confirming FN). Erroneous extractions were captured as edge cases to iteratively refine retrieval parameters, prompt logic, and conflict-resolution rules. This process continued until deployment thresholds of ≥95% accuracy and ≥95% precision (Positive Predictive Value, PPV) were met.

#### Reported Performance Metrics

Reviewers classified each field of extracted data, and we derived True Positive (TP), True Negative (TN), False Positive (FP), or False Negative (FN) from which the following metrics were calculated:

- **Accuracy** = (TP + TN) / (TP + TN + FP + FN)
- **Precision (PPV)** = TP / (TP + FP)
- **Recall (Sensitivity)** = TP / (TP + FN)
- **F1 Score** = 2 × (Precision × Recall) / (Precision + Recall)

Extraction models undergo continuous review, version-controlled updates, and consistency checks, with version control enabling rollback.

## Results

Medical records in C-CDA and image formats were utilized for LLM extractor development and deployment according to the methods herein (Figure 1). Three schemas were designed to extract medications, radiation therapy procedures, and surgical procedures from clinical documents. Medication schema elements included the medication name, start and end dates, indication, discontinuation reason, dose, and route of administration. Radiation therapy schema elements included the type of radiation, start and end dates, planned doses and fractions, and received dosage. Surgical procedure schema elements included the procedure description, date, anatomic site, reason, extent of resection, and histopathology findings.

**Figure 1.**
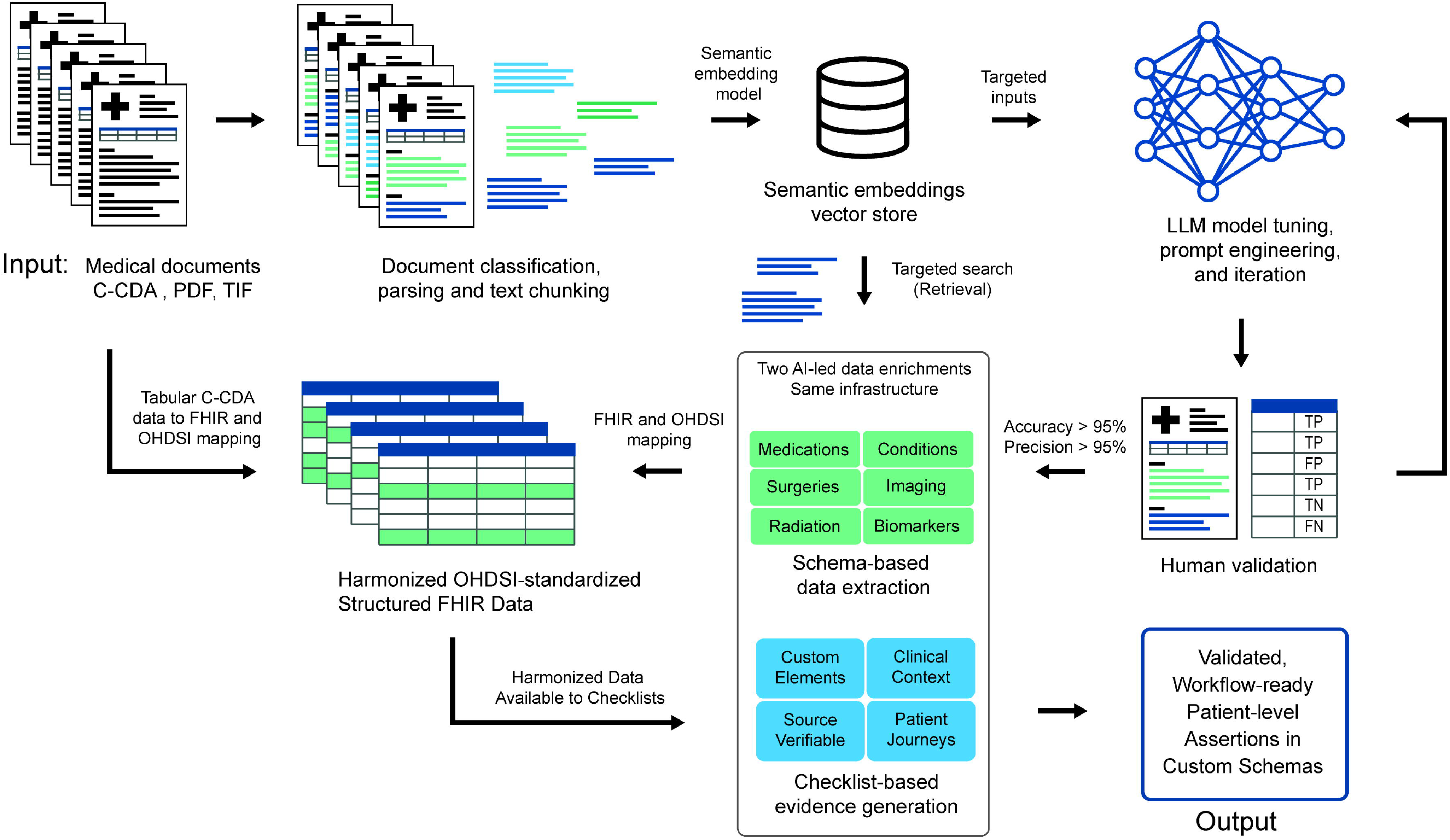
Extraction pipeline for development, validation, and deployment of LLMs. Patient documents are received in multiple formats, including C-CDA, FHIR, and image-based files, requiring preprocessing steps such as document parsing, OCR, and structured metadata extraction for interoperability. Text is segmented into chunks using section-based and semantic chunking strategies, followed by embedding-based classification. Targeted semantic search and document classification algorithms are utilized for retrieval of appropriate documents for LLM model development and deployment. Performance metric thresholds are utilized to graduate LLMs from development to human validation to large-scale deployment. LLM-extracted data is mapped to FHIR R4 resources, normalized to OMOP concepts, and integrated with tabular C-CDA data ensuring standardized representation and enhancing data completeness. In parallel, a retrieval-augmented checklist system interrogates embedded document text to extract patient-level clinical assertions across the longitudinal record. Outputs are generated as structured variables accompanied by supporting evidence and source-document citations.

For medication extraction, a cohort of 38 patients was selected for initial validation. An embeddings-based search of terms such as “chemotherapy,” “immunotherapy,” and “medications” yielded 216 medical documents (median 5 [IQR 4–9] per patient) on which to deploy the extraction process. A total of 5,789 medications were extracted, yielding 3,002 distinct medication records after removing duplicates. Radiation procedure and surgical procedure extraction were validated on 40 patients (408 documents; median 19 [IQR 10–23]) and 34 patients (108 documents; median 4 [IQR 2–7]), respectively. Extracted data elements were manually validated against source records to calculate accuracy, precision, recall, and F1 score. Overall performance was consistent across all three schemas, reaching ∼95% or greater performance across all measures and data fields (Table 2).

**Table 2.**
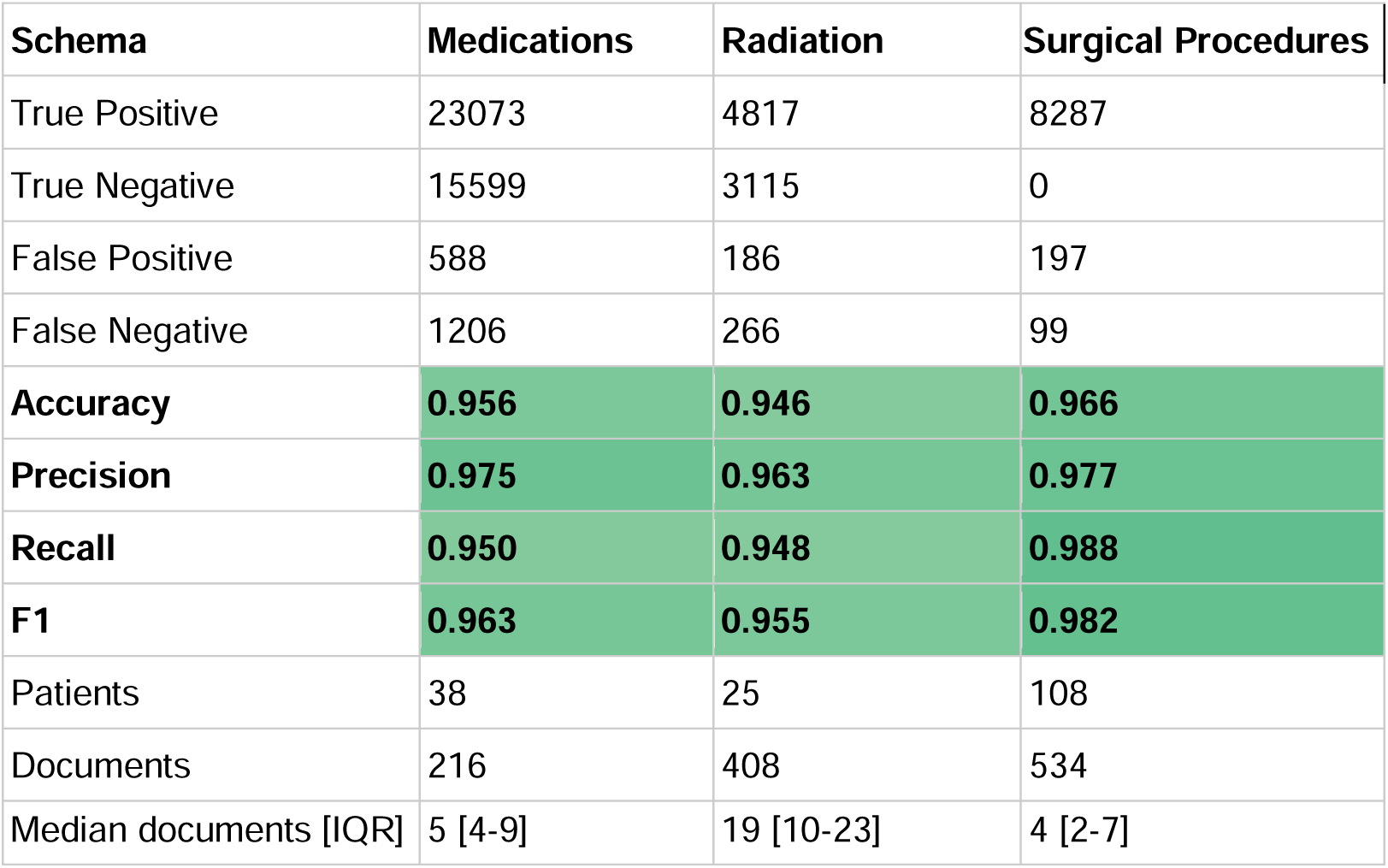
Performance metrics of LLM schema-based extractors.

The LLM extractors were then deployed to the medical records of 3,493 patients with anticipated cancer diagnoses and compared with data available pre-structured from C-CDA (Table 3). A significant amount of medication data was available for these patients in structured format after conversion from C-CDA to FHIR, yielding over 2.25 million individual medication records and 1,418 distinct drug ingredients from 104,348 medical documents. Separately, LLM-based extraction yielded ∼1.6 million medication records and 2,281 distinct drug ingredients from 92,914 medical documents after normalization to OHDSI vocabularies.

**Table 3.**
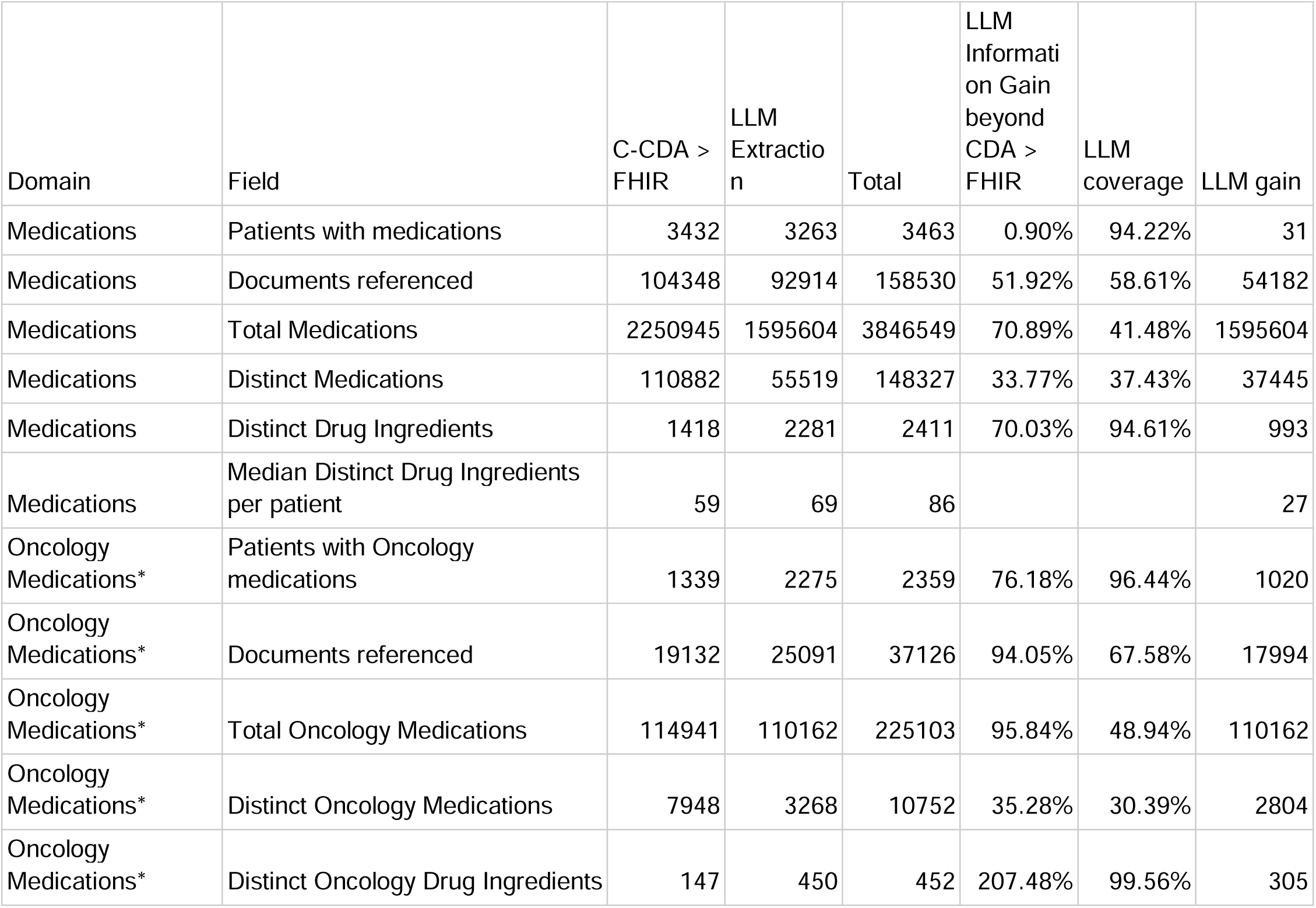
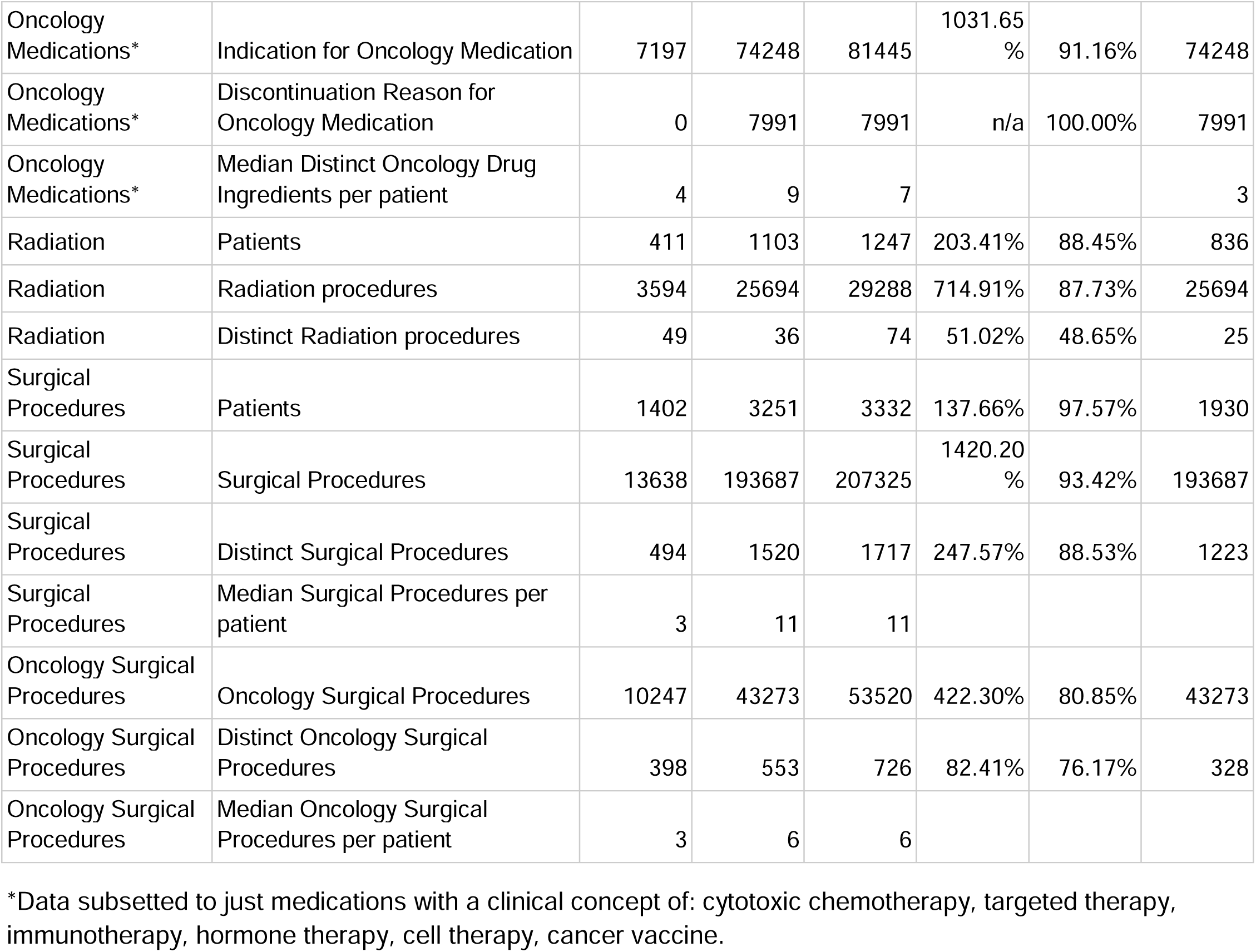
Information gain from deployed LLM extractors.

| Domain | Field | C-CDA > FHIR | LLM Extraction | Total | LLM Information Gain beyond CDA > FHIR | LLM coverage | LLM gain |
| --- | --- | --- | --- | --- | --- | --- | --- |
| Medications | Patients with medications | 3432 | 3263 | 3463 | 0.90% | 94.22% | 31 |
| Medications | Documents referenced | 104348 | 92914 | 158530 | 51.92% | 58.61% | 54182 |
| Medications | Total Medications | 2250945 | 1595604 | 3846549 | 70.89% | 41.48% | 1595604 |
| Medications | Distinct Medications | 110882 | 55519 | 148327 | 33.77% | 37.43% | 37445 |
| Medications | Distinct Drug Ingredients | 1418 | 2281 | 2411 | 70.03% | 94.61% | 993 |
| Medications | Median Distinct Drug Ingredients per patient | 59 | 69 | 86 |  |  | 27 |
| Oncology Medications* | Patients with Oncology medications | 1339 | 2275 | 2359 | 76.18% | 96.44% | 1020 |
| Oncology Medications* | Documents referenced | 19132 | 25091 | 37126 | 94.05% | 67.58% | 17994 |
| Oncology Medications* | Total Oncology Medications | 114941 | 110162 | 225103 | 95.84% | 48.94% | 110162 |
| Oncology Medications* | Distinct Oncology Medications | 7948 | 3268 | 10752 | 35.28% | 30.39% | 2804 |
| Oncology Medications* | Distinct Oncology Drug Ingredients | 147 | 450 | 452 | 207.48% | 99.56% | 305 |
| Oncology Medications* | Indication for Oncology Medication | 7197 | 74248 | 81445 | 1031.65 % | 91.16% | 74248 |
| Oncology Medications* | Discontinuation Reason for Oncology Medication | 0 | 7991 | 7991 | n/a | 100.00% | 7991 |
| Oncology Medications* | Median Distinct Oncology Drug Ingredients per patient | 4 | 9 | 7 |  |  | 3 |
| Radiation | Patients | 411 | 1103 | 1247 | 203.41% | 88.45% | 836 |
| Radiation | Radiation procedures | 3594 | 25694 | 29288 | 714.91% | 87.73% | 25694 |
| Radiation | Distinct Radiation procedures | 49 | 36 | 74 | 51.02% | 48.65% | 25 |
| Surgical Procedures | Patients | 1402 | 3251 | 3332 | 137.66% | 97.57% | 1930 |
| Surgical Procedures | Surgical Procedures | 13638 | 193687 | 207325 | 1420.20 % | 93.42% | 193687 |
| Surgical Procedures | Distinct Surgical Procedures | 494 | 1520 | 1717 | 247.57% | 88.53% | 1223 |
| Surgical Procedures | Median Surgical Procedures per patient | 3 | 11 | 11 |  |  |  |
| Oncology Surgical Procedures | Oncology Surgical Procedures | 10247 | 43273 | 53520 | 422.30% | 80.85% | 43273 |
| Oncology Surgical Procedures | Distinct Oncology Surgical Procedures | 398 | 553 | 726 | 82.41% | 76.17% | 328 |
| Oncology Surgical Procedures | Median Oncology Surgical Procedures per patient | 3 | 6 | 6 |  |  |  |

\*Data subsetted to just medications with a clinical concept of: cytotoxic chemotherapy, targeted therapy, immunotherapy, hormone therapy, cell therapy, cancer vaccine.

Document selection for the LLM-based extraction identified 54,182 additional medical documents for data extraction beyond those containing pre-structured medications (52% more), and generated an increase in 71% more total medications and 70% more distinct drug ingredients than what was available in the C-CDA medication tables. Importantly, a review of oncology-specific medications showed that only 1,399 patients (40%) had a structured oncology medication in their medical record. The LLM extracted 110,162 oncology medication and 452 distinct oncology drug ingredients from 25,091 medical documents (of which 71% did not contain pre-structured medications) across 2,275 patients (65%). This yielded a 76% increase in the number of patients with at least one oncology medication, a 96% increase in total oncology medication records, and a 207% increase in distinct oncology drug ingredients. The LLM extracted 305 unique oncology therapies/medications not found in any structured C-CDA table. The LLM also extracted enhanced medication variables, outperforming the pre-structured C-CDA oncology medication data for the indication for prescription (70.5% vs. 4.6%) and reason for medication discontinuation (9.7% vs. 0%).

In contrast to medications, radiation and surgical procedure data were largely absent from pre-structured C-CDA records, making LLM extraction the primary source of structured data for these domains. For radiation, C-CDA yielded only 3,594 procedures across 411 patients, while LLM extraction expanded coverage to 25,694 procedures across 1,103 patients — a 715% increase in total procedures and providing 88% coverage of all radiation procedures. For surgical procedures, LLM extraction similarly transformed coverage: compared with 13,638 procedures across 1,402 patients from C-CDA, LLM extraction yielded 193,687 total and 1,520 distinct procedures across 3,351 patients, including a 422% increase in oncology-specific surgical records.

We next designed checklist-based data extraction systems to generate (a) the initial and current diagnosis details, and (b) oncology lines of therapy and outcomes. Diagnosis schemas included the cancer type (e.g. breast, lung, colorectal), diagnosis, histology, initial diagnosis date or most recent staging date, stage, TNM stage, grade, tumor size, metastatic sites, and a source flag to define whether the primary evidence was extracted from pathology, clinic note, or imaging records. The lines of therapy checklist leveraged anti-cancer treatments extracted by the schema-based extractors (medications and radiation therapy) and clinical context from oncology progress notes and imaging reports to assert the line of therapy, compile regimens with start and end dates, define the best response (complete response, partial response, stable disease, progressive disease) and best response date, the reason for regimen discontinuation, the date of progression (if defined), and the evidence level for progression (radiological or clinical). Checklists were deployed against 99 patients with confirmed cancer diagnoses and 3,960 discrete elements were manually reviewed according to the Methods. Together, the diagnosis checklist reached 98.2% accuracy, 98.7% precision, 99.4% recall, and 99.0% F1, and the lines of therapy checklist reached 97.0% accuracy, 95.4% precision, 99.8% recall, and 97.6% F1 (Table 4). The median number of total documents per patient was 209.5 but the median number of documents cited per diagnosis checklist item was four and per lines of therapy checklist item was eight, indicating that semantic retrieval substantially reduced the document burden for both extraction and reviewer verification.

**Table 4.** Performance Metrics of Diagnosis Details and Lines of Therapy Checklists.

| **Checklist** | **Cancer**<br>**Diagnosis and**<br>**Tumor**<br>**Characteristic**<br>**s** | **Lines of**<br>**Therapy and**<br>**Outcomes** |
| --- | --- | --- |
| Total elements reviewed | 3960 | 842 |
| True Positive | 3642 | 501 |
| True Negative | 246 | 316 |
| False Positive | 49 | 24 |
| False Negative | 23 | 1 |
| **Accuracy** | **0.982** | **0.970** |
| **Precision (PPV)** | **0.987** | **0.954** |
| **Recall (Sensitivity)** | **0.994** | **0.998** |
| **F1** | **0.990** | **0.976** |
| Median total documents | 209.5 | 273 |
| IQR total documents | 106-425 | 135-483 |
| Median documents referenced by checklist | 4 | 8 |

Deploying these validated checklists against the same 3,493 patient cohort yielded 3,247 (93.5%) with initial diagnosis and diagnosis date. Cancer stage was extracted for 64% of patients, grade for 62%, and TNM for 50%+ of patients. These data were in marked contrast to structured cancer diagnoses with an onset date, available for 2,437 patients (69.8%), where condition names only occasionally contained stage references such as “metastatic” (228 patients, 6.5%) and rarely contained grade information such as “poorly differentiated” (17 patients, 0.5%). The checklist flagged pathology as the primary source document used for 83.6% of patients, providing further confidence in the extracted data. The lines of therapy checklist, which referenced the medication and radiation data, yielded 2,320 patients with at least one line extracted (66.4%) and 4,218 total lines of therapy. First-line therapy constituted the most prevalent treatment (43.17%), followed by second-line (15.15%), adjuvant (9.67%), third-line (6.02%), neoadjuvant (4.20%), and fourth-line (2.58%) therapies within the dataset. Best response and date was available for 782 (19%) lines and Reason for discontinuation was available for 894 (21%) lines. Among reason for discontinuation, Completion of Planned Therapy was most common (57%), followed by Adverse Events or Intolerance (19%), Disease Progression (15%), Patient Preference (7%), and Provider Changed Treatment Plan (2%). Dates of progression were available for 280 lines of therapy (7% of all lines). An example of synthesizing all of these endpoints to provide a graphical longitudinal journey for one patient is presented in Figure 2.

**Figure 2.**
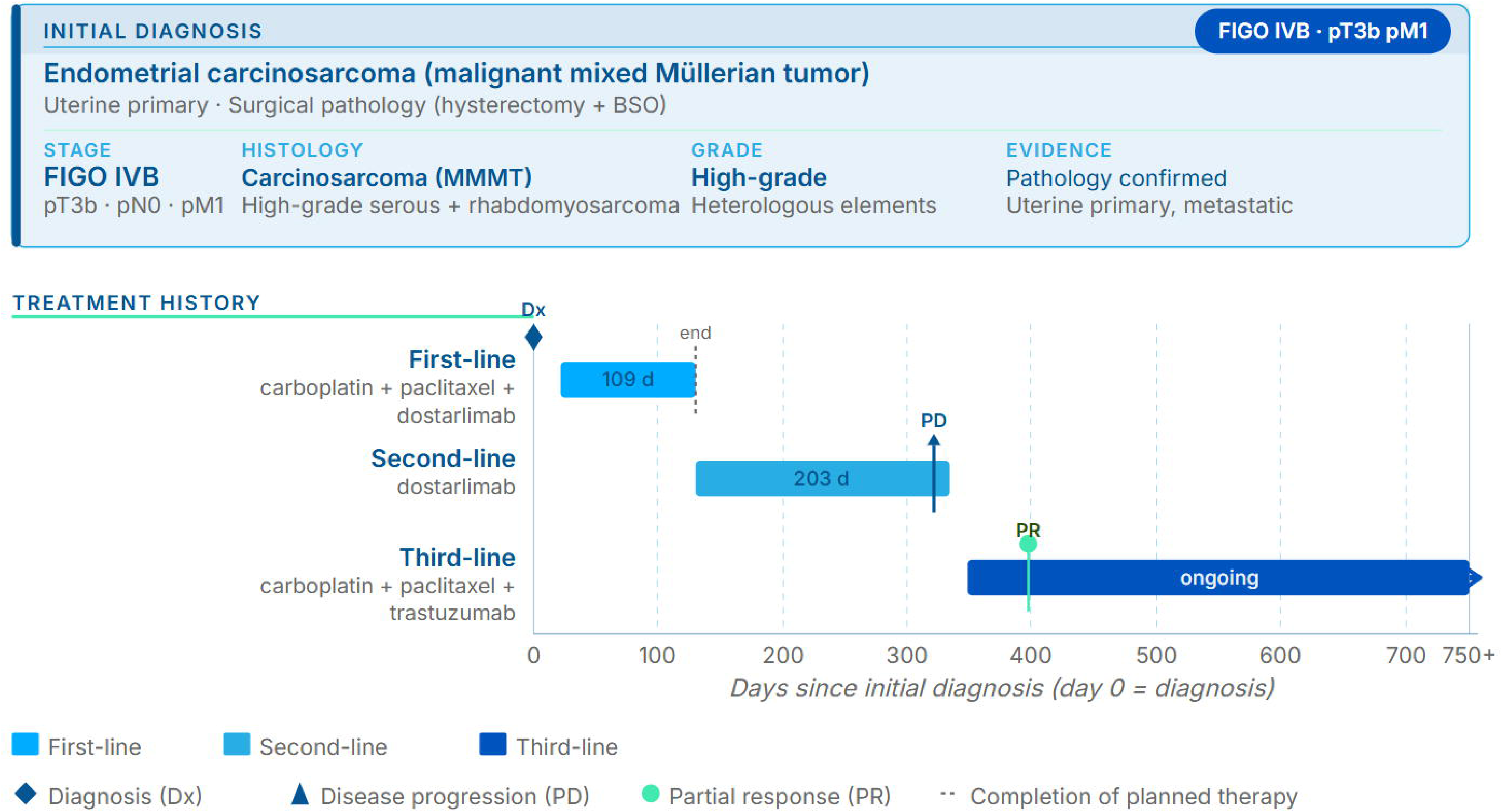
Sample patient treatment journey from integrated LLM extractions and checklist results. Data leveraging schema-based LLMs for medications and surgical procedures and checklist results for diagnosis details and lines of therapy were utilized to generate a graphical summary for a single patient. Day 0 was set at the Initial Diagnosis Date element, and other dates were used relationally.

To characterize the analytical utility of the extracted longitudinal data, we examined treatment patterns and response outcomes across the 3,493-patient cohort. Among patients with at least one line of therapy extracted, the cohort represented a heterogeneous mix of solid tumor types, with lung cancer (n=1,068), breast cancer (n=1,035), and colorectal cancer (n=407) comprising the largest subgroups (Figure 3A). Time on treatment varied across lines of therapy: median treatment duration was 91 days at first line, 90 days at second line, 133 days at third line, and 96 days at fourth line, with wide interquartile ranges at each line reflecting the heterogeneity of treatment regimens and patient populations (Figure 3B). Radiation-only episodes were excluded from this analysis. Median first-line treatment duration assessed by cancer type ranged from 37 days for esophageal cancer (n=35) to 147 days for breast cancer (n=85), with lung (63 days, n=155), colorectal (112 days, n=39), endometrial (105 days, n=19), and ovarian (130 days, n=27) cancers spanning the intermediate range (Figure 3C). The most prominent first-line therapeutic regimens and interventions are provided in Figure 3D. Among patients with a documented best response, objective response rate (ORR; complete or partial response) was 64.7% (95% CI 59.0–70.0%, n=286) at first line, declining to 36.2% (95% CI 28.4–44.9%, n=127) at second line and 21.5% (95% CI 13.3–33.0%, n=65) at third line. Disease control rate (DCR; complete response, partial response, or stable disease) followed a similar pattern, from 84.6% (95% CI 80.0–88.3%) at first line to 53.8% (95% CI 41.9–65.4%) at third line (Figure 4A). First-line ORR varied across tumor types, from 42.9% (95% CI 24.5–63.5%, n=21) in colorectal cancer to 85.0% (95% CI 64.0–94.8%, n=20) in esophageal cancer, though confidence intervals were wide for several subgroups given limited sample sizes (Figure 4B).

**Figure 3.**
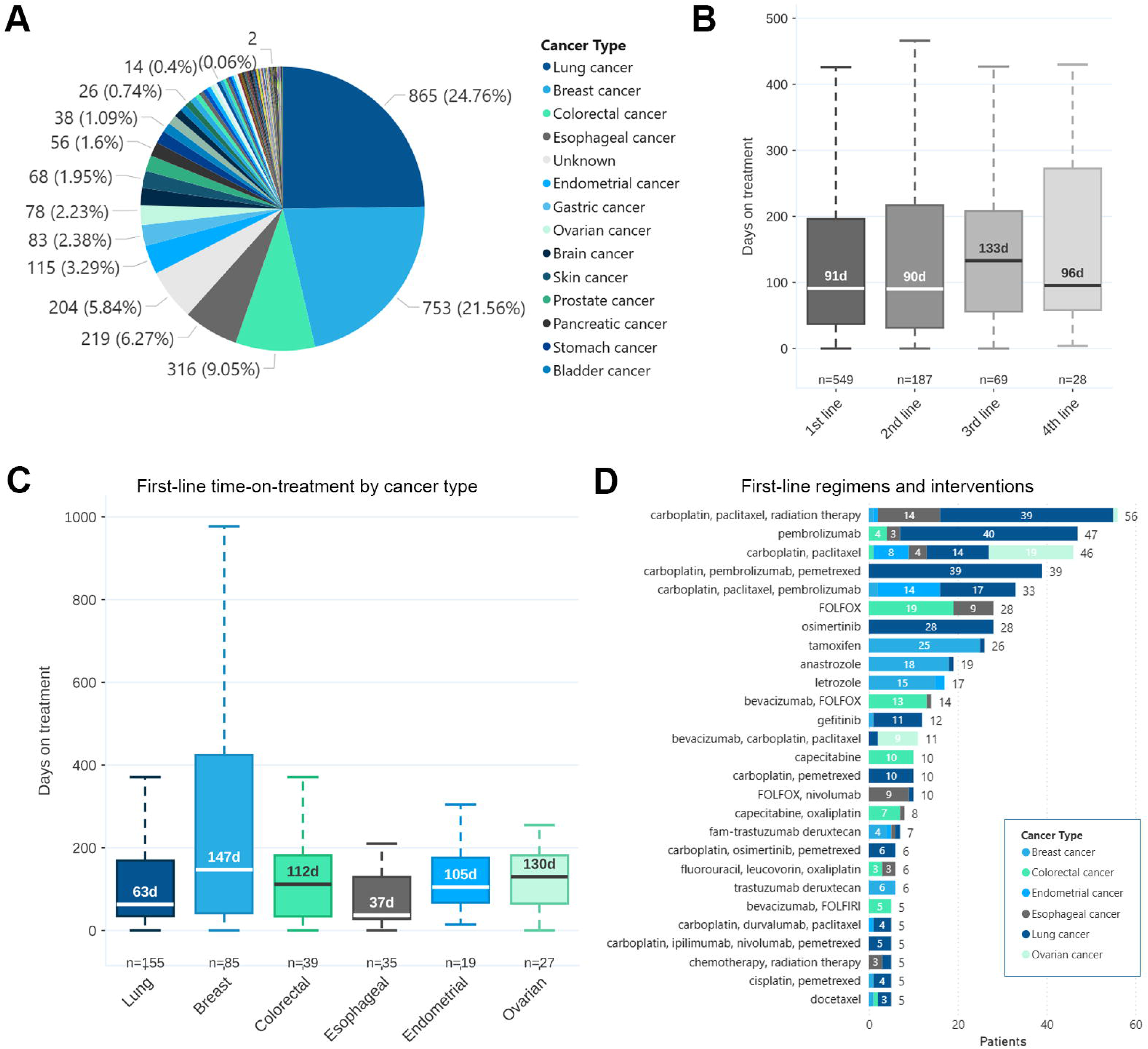
Cohort level integration of Diagnosis Details and Lines of Therapy checklist results. (A) Distribution of patients by cancer type, as assessed by the diagnosis details checklist. (B) Distribution of days on treatment across sequential lines of therapy, excluding radiation-only episodes. Sample sizes reflect unique treatment episodes with documented treatment duration. (C) Distribution of days on first-line treatment for the six most common cancer types in the cohort, excluding radiation-only episodes. Cancer types are ordered left to right by decreasing cohort frequency. (D) The most frequent regimens or interventions for first-line treatment among the top cancer types. In (B) and (C), the box represents the interquartile range (IQR), the horizontal line indicates the median, and whiskers extend to 1.5×IQR. Median values are labeled within or above each box. Outliers beyond 1.5×IQR are not displayed.

**Figure 4.**
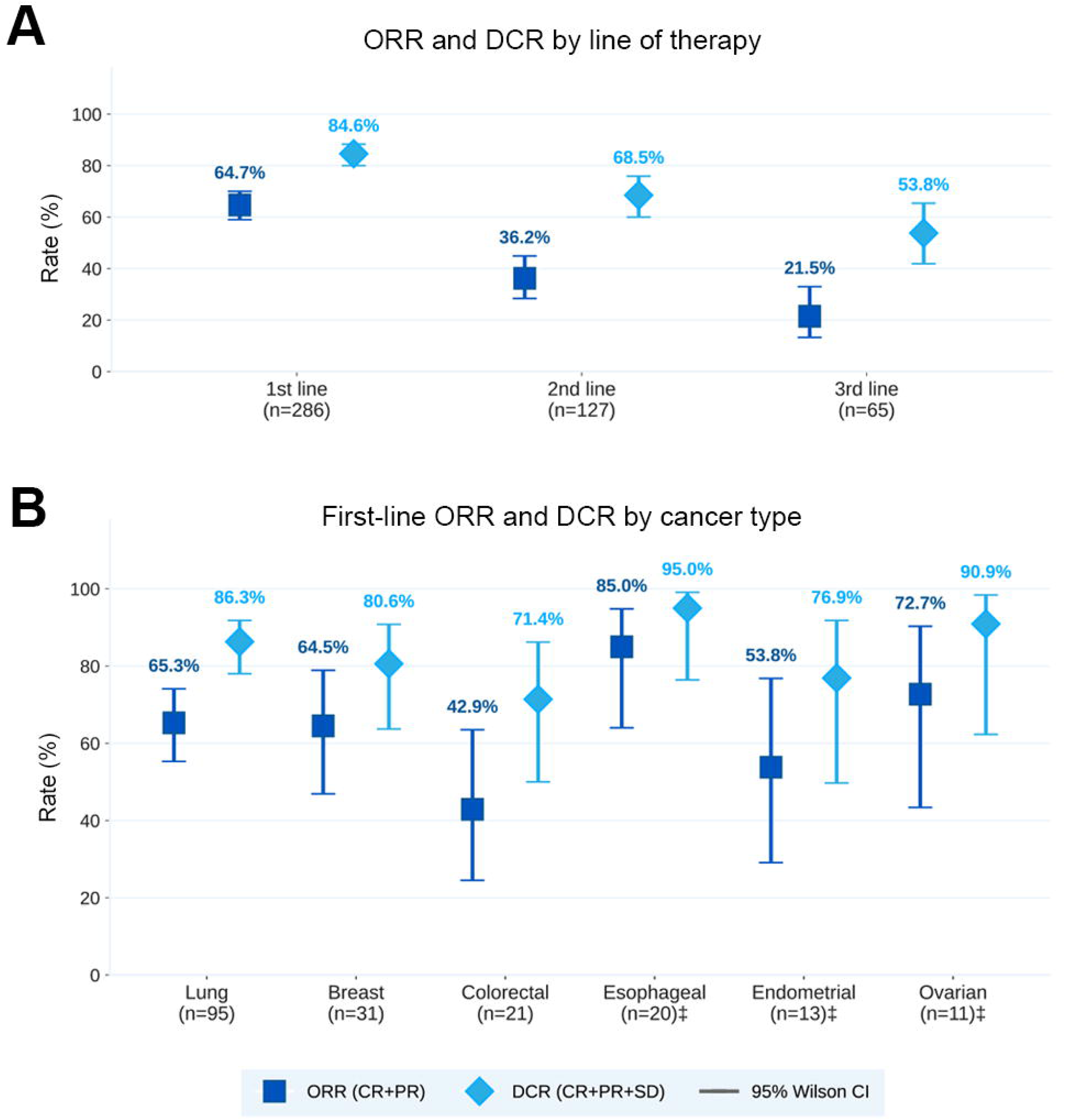
Objective response rate and disease control rate by line of therapy and cancer type. Point estimates and 95% Wilson score confidence intervals for objective response rate (ORR; CR+PR, filled squares) and disease control rate (DCR; CR+PR+SD, filled diamonds) among response-evaluable patients with documented best response. (A) ORR and DCR across first, second, and third lines of therapy. (B) First-line ORR and DCR stratified by cancer type. Response reflects semantics present in medical records from treating clinicians and radiologists; independent radiological review was not performed. ‡ Denotes groups with n<25, where confidence intervals are wide and point estimates should be interpreted with caution.

## Discussion

The analyses presented here illustrate how structured, longitudinal oncology data — encompassing diagnosis, treatment sequencing, response, and outcomes — can be systematically generated from real-world medical records at scale. This single deployment of the platform yielded analysis-ready cohort-level data across thousands of patients, spanning multiple tumor types and lines of therapy, underscoring the practical utility of this approach for data extraction and evidence generation.

Our findings demonstrate the effectiveness of LLM-based extraction for retrieving and structuring medication data from diverse medical record formats, significantly augmenting structured EHR data with important clinical information. The LLM-based approach increased both the total number of medication elements structured and the distinct number of drug ingredients extracted, particularly enhancing oncology-specific medication retrieval. This highlights the potential for LLMs to bridge gaps in structured clinical documentation by extracting clinically relevant information from unstructured or semi-structured sources.

In addition to schema-based extraction, the checklist framework demonstrated that LLMs can reliably generate patient-level clinical assertions across longitudinal medical records. By combining semantic retrieval with prescriptive prompts, the system identified and reconciled clinically relevant information distributed across large document collections — a median of 209.5 documents per patient — while reducing the evidence burden to a median of four cited documents per diagnosis item and eight per lines-of-therapy item. Critically, the two extraction methods are deeply integrated: schema-based medication and radiation extraction feeds directly into the lines-of-therapy checklist, enabling the system to assert treatment sequences, regimen dates, best response, and discontinuation reasons as coherent patient-level clinical summaries rather than isolated data points. This layered architecture — from granular document-level event extraction to patient-level assertion — is reproducible for a diversity of clinical abstraction workflows, enabling scalable extraction of complex variables beyond oncology diagnosis and treatment details.

Our approach achieved high F1 scores across multiple extracted data elements (average ∼95%). However, some limitations warrant consideration. The accuracy of LLM-based extraction is contingent on high-quality input documents, and variations in OCR performance for scanned documents, such as faxes embedded within the medical record, may introduce errors or omissions. Additionally, while LLMs exhibited strong performance in the NER and RE tasks (identifying and correctly associating a dose, route, etc. with a specific medication), strict validation steps were necessary to mitigate hallucinations and ensure that only explicitly stated extractions were retained. While developed with a focus on F1 scores, we note that different applications may require optimization for one metric over another.

Careful prompt engineering with version control remains a critical step to ensure the fidelity of LLM output. Schemas should be optimized for specific tasks and document types for reliable clinical data extraction that integrates with existing coding systems. This highlights the importance of maintaining interoperability through mappings to standard terminologies like FHIR and OMOP. Our approach ensured that extracted data aligned with established clinical dictionaries, reducing the risk of data fragmentation or misclassification.

Future work will explore fine-tuning LLMs for domain-specific extractions, refinement of document chunking strategies and text classifications, extension of schema elements, and adopting additional validation metrics. For example, our current validation is heavily biased against the LLM, with two human abstractors and a referee evaluating the LLM results. Double data entry with a referee was a common approach for validation of data entry in clinical trials, with a mean reported error rate of 0.14%.^12^ While this method establishes a gold standard for measuring LLM performance, in future work it may be useful to establish inter-rater reliability between human data abstractors in order to position LLM extraction relative to expert human performance.

Despite these additional considerations, our results provide compelling evidence that LLM-based extraction can meaningfully enhance clinical data accessibility and completeness. The methodology presented here — combining schema-based extraction, checklist-driven patient-level assertion, and semantic retrieval across heterogeneous medical records — establishes a scalable and validated approach to generating interoperable, analysis-ready clinical datasets. As the volume and complexity of real-world healthcare data continue to grow, platforms capable of transforming fragmented records into structured, traceable, and clinically meaningful outputs will be essential for enabling precision oncology research, supporting clinical trial matching, and accelerating evidence-based care.

## Data Availability

Data produced in the present study are available upon reasonable request to the authors.

## Acknowledgments

We would like to acknowledge Sabrina Irizarry, Hannah Hayes, Jacob Hendersot, Zachary Kaufman, and Joshua Marker.

