## Supplemental Information for "A Scalable Method for Validated Data Extraction from Electronic Health Records with Large Language Models"

**Supplemental Table 1: Full schema-based LLM extractor performance metrics**

| **Schema** | **Field** | **True Positive** | **True Negative** | **False Positive** | **False Negative** | **Accuracy** | **Precision** | **Recall** | **F1** |
| --- | --- | --- | --- | --- | --- | --- | --- | --- | --- |
| Medications | medication_name | 5779 | 2 | 5 | 0 | 0.9991 | 0.9991 | 1.0000 | 0.9996 |
| Medications | start_date | 3312 | 2027 | 101 | 345 | 0.9229 | 0.9704 | 0.9057 | 0.9369 |
| Medications | end_date | 1469 | 3955 | 74 | 286 | 0.9378 | 0.9520 | 0.8370 | 0.8908 |
| Medications | indication | 2771 | 2521 | 229 | 266 | 0.9145 | 0.9237 | 0.9124 | 0.9180 |
| Medications | discontinuation_reason | 156 | 5572 | 16 | 15 | 0.9946 | 0.9070 | 0.9123 | 0.9096 |
| Medications | dose | 4746 | 792 | 101 | 139 | 0.9585 | 0.9792 | 0.9715 | 0.9753 |
| Medications | route | 4840 | 730 | 62 | 155 | 0.9625 | 0.9874 | 0.9690 | 0.9781 |
| **Medications** | **Overall** | **23073** | **15599** | **588** | **1206** | **0.9557** | **0.9751** | **0.9503** | **0.9626** |
| Radiation | procedure_name | 697 | 0 | 2 | 0 | 0.9971 | 0.9971 | 1.0000 | 0.9986 |
| Radiation | status | 600 | 51 | 22 | 25 | 0.9327 | 0.9646 | 0.9600 | 0.9623 |
| Radiation | discontinuation_reason | 494 | 174 | 8 | 22 | 0.9570 | 0.9841 | 0.9574 | 0.9705 |
| Radiation | description | 655 | 0 | 44 | 0 | 0.9371 | 0.9371 | 1.0000 | 0.9675 |
| Radiation | start_date | 382 | 262 | 30 | 25 | 0.9213 | 0.9272 | 0.9386 | 0.9328 |
| Radiation | end_date | 410 | 217 | 36 | 35 | 0.8983 | 0.9193 | 0.9213 | 0.9203 |
| Radiation | fractions_planned | 217 | 436 | 12 | 34 | 0.9342 | 0.9476 | 0.8645 | 0.9042 |
| Radiation | fractions_completed | 188 | 464 | 12 | 35 | 0.9328 | 0.9400 | 0.8430 | 0.8889 |
| Radiation | total_dose_planned | 172 | 494 | 4 | 29 | 0.9528 | 0.9773 | 0.8557 | 0.9125 |
| Radiation | total_dose_completed | 150 | 516 | 4 | 29 | 0.9528 | 0.9740 | 0.8380 | 0.9009 |
| Radiation | dosage_units | 175 | 496 | 1 | 27 | 0.9599 | 0.9943 | 0.8663 | 0.9259 |
| Radiation | anatomic_site | 677 | 5 | 11 | 5 | 0.9771 | 0.9840 | 0.9927 | 0.9883 |
| **Radiation** | **Overall** | **4817** | **3115** | **186** | **266** | **0.9461** | **0.9628** | **0.9477** | **0.9552** |
| Surgical Procedures | description | 1428 | 0 | 13 | 0 | 0.991 | 0.991 | 1.000 | 0.995 |
| Surgical Procedures | date | 1368 | 0 | 50 | 10 | 0.958 | 0.965 | 0.993 | 0.979 |
| Surgical Procedures | anatomic_site | 1402 | 0 | 19 | 7 | 0.982 | 0.987 | 0.995 | 0.991 |
| Surgical Procedures | reason | 1341 | 0 | 64 | 24 | 0.938 | 0.954 | 0.982 | 0.968 |
| Surgical Procedures | extent_of_resection | 1388 | 0 | 27 | 12 | 0.973 | 0.981 | 0.991 | 0.986 |
| Surgical Procedures | histopathology_findings | 1360 | 0 | 24 | 46 | 0.951 | 0.983 | 0.967 | 0.975 |
| **Surgical Procedures** | **Overall** | **8287** | **0** | **197** | **99** | **0.966** | **0.977** | **0.988** | **0.982** |

**Supplemental Table 2: Full validation performance metrics for Cancer Diagnosis and Tumor Characteristics Checklist**

| Checklist Item | Property | True Positive | True Negative | False Positive | False Negative | Accuracy | Precision (PPV) | Recall (Sensitivity) | F1 |
| --- | --- | --- | --- | --- | --- | --- | --- | --- | --- |
| Initial Diagnosis Details | Cancer Type | 99 | 0 | 0 | 0 | 1.000 | 1.000 | 1.000 | 1.000 |
| Initial Diagnosis Details | Initial Diagnosis | 99 | 0 | 0 | 0 | 1.000 | 1.000 | 1.000 | 1.000 |
| Initial Diagnosis Details | Initial Histology | 99 | 0 | 0 | 0 | 1.000 | 1.000 | 1.000 | 1.000 |
| Initial Diagnosis Details | Initial Diagnosis Date | 93 | 0 | 6 | 0 | 0.939 | 0.939 | 1.000 | 0.969 |
| Initial Diagnosis Details | Initial Cancer Stage | 74 | 23 | 0 | 2 | 0.980 | 1.000 | 0.974 | 0.987 |
| Initial Diagnosis Details | Initial TNM | 78 | 20 | 1 | 0 | 0.990 | 0.987 | 1.000 | 0.994 |
| Initial Diagnosis Details | Initial Grade | 77 | 20 | 1 | 1 | 0.980 | 0.987 | 0.987 | 0.987 |
| Initial Diagnosis Details | Evidence | 99 | 0 | 0 | 0 | 1.000 | 1.000 | 1.000 | 1.000 |
| **Initial Diagnosis Details** | **Overall** | **619** | **63** | **8** | **3** | **0.984** | **0.987** | **0.995** | **0.991** |
| Current Diagnosis Details | Cancer Type | 99 | 0 | 0 | 0 | 1.000 | 1.000 | 1.000 | 1.000 |
| Current Diagnosis Details | Current Diagnosis | 99 | 0 | 0 | 0 | 1.000 | 1.000 | 1.000 | 1.000 |
| Current Diagnosis Details | Current Histology | 99 | 0 | 0 | 0 | 1.000 | 1.000 | 1.000 | 1.000 |
| Current Diagnosis Details | Current Staging Date | 89 | 0 | 10 | 0 | 0.899 | 0.899 | 1.000 | 0.947 |
| Current Diagnosis Details | Current Cancer Stage | 78 | 19 | 1 | 1 | 0.980 | 0.987 | 0.987 | 0.987 |
| Current Diagnosis Details | TNM | 82 | 14 | 2 | 1 | 0.970 | 0.976 | 0.988 | 0.982 |
| Current Diagnosis Details | Current Cancer Grade | 70 | 27 | 1 | 1 | 0.980 | 0.986 | 0.986 | 0.986 |
| Current Diagnosis Details | Evidence | 95 | 0 | 3 | 1 | 0.960 | 0.969 | 0.990 | 0.979 |
| **Current Diagnosis Details** | **Overall** | **616** | **60** | **14** | **3** | **0.975** | **0.978** | **0.995** | **0.986** |
| Tumor Details | Lymph Nodes Tested | 93 | 0 | 1 | 5 | 0.939 | 0.989 | 0.949 | 0.969 |
| Tumor Details | Node Status | 99 | 0 | 0 | 0 | 1.000 | 1.000 | 1.000 | 1.000 |
| Tumor Details | Metastatic Sites | 99 | 0 | 0 | 0 | 1.000 | 1.000 | 1.000 | 1.000 |
| Tumor Details | Tumor Size | 99 | 0 | 0 | 0 | 1.000 | 1.000 | 1.000 | 1.000 |
| Tumor Details | Tumor Laterality | 99 | 0 | 0 | 0 | 1.000 | 1.000 | 1.000 | 1.000 |
| **Tumor Details** | **Overall** | **489** | **0** | **1** | **5** | **0.988** | **0.998** | **0.990** | **0.994** |
| **Full Diagnosis Details Checklist** | **Overall** | **3642** | **246** | **49** | **23** | **0.982** | **0.987** | **0.994** | **0.99** |

**Supplemental Table 3: Full validation performance metrics for Lines of Therapy and Outcomes Checklist**

|  | **True Positive** | **True Negative** | **False Positive** | **False Negative** | **Accuracy** | **Precision (PPV)** | **Recall (Sensitivity)** | **F1** |
| --- | --- | --- | --- | --- | --- | --- | --- | --- |
| Line of Therapy | 77 | 8 | 0 | 0 | 1.000 | 1.000 | 1.000 | 1.000 |
| Regimen or Interventions | 76 | 8 | 0 | 1 | 0.988 | 1.000 | 0.987 | 0.993 |
| Start Date | 66 | 9 | 6 | 0 | 0.926 | 0.917 | 1.000 | 0.957 |
| End Date | 62 | 13 | 6 | 0 | 0.926 | 0.912 | 1.000 | 0.954 |
| Best Response | 27 | 54 | 4 | 0 | 0.953 | 0.871 | 1.000 | 0.931 |
| Best Response Date | 25 | 55 | 5 | 0 | 0.941 | 0.833 | 1.000 | 0.909 |
| Reason for Discontinuation | 58 | 26 | 1 | 0 | 0.988 | 0.983 | 1.000 | 0.991 |
| Standardized Reason for Discontinuation | 72 | 13 | 0 | 0 | 1.000 | 1.000 | 1.000 | 1.000 |
| Date of Progression | 22 | 61 | 2 | 0 | 0.976 | 0.917 | 1.000 | 0.957 |
| Progression Evidence Level | 16 | 69 | 0 | 0 | 1.000 | 1.000 | 1.000 | 1.000 |
| Overall | 501 | 316 | 24 | 1 | 0.970 | 0.954 | 0.998 | 0.976 |
